# The impact of COVID-19 Pandemic on Domestic Gender Based Violence Against Female HIV and Tuberculosis Patients in Timor-Leste: A Qualitative Study

**DOI:** 10.1101/2023.08.28.23294701

**Authors:** Nelson Martins, Domingos Soares, Caetano Gusmao, Maria Nunes, Laura Abrantes, Diana Valadares, Suzi Marcal, Marcelo Mali, Luis Alves, Jorge Martins, Valente da Silva

**Affiliations:** Daslo Research and Development; Instituto Nacional de Saúde Publica Timor-Leste (INSP-TL), Ministry of Health Timor-Leste; Universidade da Paz (UNPAZ); Menzies school of Health Research, Darwin, NT-Australia; Universitas Airlangga (UNAIR), Fakultas Keperawatan; Ministerio da Saúde (MdS) Timor-Leste

**Keywords:** GBV, Gender, Violence, HIV, tuberculosis, COVID-19 pandemic

## Abstract

**Background:** Globally, there is still widespread of violence against women and girls. Timor Leste reports high prevalence of gender-based violence with 30% women have experienced intimate partner (IPV) or non-partner sexual violence. Several studies reported about the increase of domestic violence gender-based violence (D-GBV) against girls and women worldwide during the COVID-19 pandemic lockdowns. To our knowledge, there are limited numbers of research examining the occurrence of D-GBV against girls and women living with HIV/AIDS and TB during the pandemic in developing countries. This study is commissioned to understand whether women living with these two diseases experienced D-GBV during the lockdowns in Timor Leste.

**Material and Methods:** This is a qualitative phenomenology study utilizing purposive sampling technic to identify and enroll participants. The study was conducted in eight municipalities from early October 2022 to end of February 2023. It considered eight independent variables – physical violence, verbal violence, phycological violence, sexual violence, sexual harassment, social-economic violence, IPV, stigma and discrimination – to identify the occurrence of D-GBV. The study included 42 in-depth interviews (IDIs) with 19 HIV participants and 23 TB participants, and three focus group discussions (FGDs). Data analysis was performed with NVIVO version 12.1 pro by identifying codes, themes, and categories.

**Result:** The D-GBV were widely reported from all municipalities. Psychological, socio-economic, verbal, and physical violence were mostly reported from participants. The COVID-19 pandemic exacerbated D-GBV, and impeded participants to apply coping mechanisms in dealing with the violence. Stigma and discrimination were prevalent in all study municipalities. The main causes of the violence were economic factors, jealousy and denial, tradition/cultural issues, and failure to perform household work. The participants reported using various coping mechanisms to deal with D-GBV, including (1) seeking external support or avoidance and (2) staying and facing the perpetrator at home. The study identified a triple vulnerability for female HIV and TB patients from experiencing various forms of D-GBV during the period of lockdown.

**Discussion and Conclusion:** While this study focused on females living with HIV/AIDS and TB only, its findings have amplified the qualitative information on the magnitude of D-GBV in Timor-Leste. The findings from this study suggest the need to pay more attention to women living with HIV and TB in order to help them to not only survive the disease(s) but also to protect them from domestic and gender-based violence. The triple vulnerabilities identified in this study reveal the weaknesses of system to combat D-GBV, stigma and discrimination against female TB and HIV patients in Timor Leste. It is recommended to train clinician working in the area of infectious diseases and reproductive health on D-GBV subject.

## Introduction

Presently, there is still widespread of violence against girls and women. It is regarded as the most devastating human rights violation, and a major impediment to the attainment of the sustainable development goals (SDGs) 2030. [1] Globally, about one in three women **(30%)** have been subjected to either physical and/or sexual intimate partner violence (IPV) or non-partner sexual violence in their lifetime. [2] Various reasons, from power imbalance, cultural, war or migrant status, trigger men to commit violence against women. [3] Situations of war, internal conflicts, natural disasters and disease pandemics that cause humanitarian crises and displacements increase violence against women, girls and children. Intimate partners and non-partner sexual violence, and new forms of violence against women are believed to worsen during humanitarian crises. [2] The health consequences of gender based violence (GBV) include risky sexual behaviour and sexually transmitted infections (STIs), physical wounds, depression, abortions, and death. [4]

The COVID-19 pandemic caused by novel coronavirus (nCoV), which emerged in China in December 2019, spread rapidly to countries around the world, and forced the World Health Organization (WHO) to declare a public health emergency, and then pandemic. [5] [6] [7] [8] [9] As of 23 September 2022, there had been 611,421,786 confirmed cases of COVID-19 globally, including 6,512,438 deaths, reported to WHO. [10] In response to the pandemic, countries introduced several public health measures to prevent and mitigate local outbreaks. These measures included country lockdowns, quarantines, stepping up surveillance, strengthening clinical and laboratory diagnostic and treatment capacities, enhancing public health interventions, and vaccination campaigns. [11] [12] [13] [14] These public health measures have been effective in controlling the transmission and reducing morbidity and mortality due to COVID-19. [11]

Unfortunately, negative consequences also emerged from the stringent measures. In particular, lockdowns interrupted routine health services and negatively impacted health gains. [15] [11] [16] The lockdowns has worsened the conditions of women living with communicable diseases (CDs) such as HIV/AIDS and tuberculosis (TB), as they prevented them to access treatment and diagnostic services. A study found that South African women with violent partners were more likely to contract HIV infection during the COVID-19 pandemic. [17] People living in small and crowded houses with poorly ventilated spaces were more exposed to COVID-19 and TB propagation. [18]

The epidemic of domestic violence against women worldwide greatly intensified during the COVID-19 pandemic. [19] The COVID-19 pandemic deepened economic and social stress, which coupled with movement restriction and social isolation measures, caused substantial increases in GBV. Sabari et al, 2020 reported about the increased global concerns on the rising numbers of domestic violence due to the direct measures to contain the spread of COVID-19. [20] The lockdowns increased the prevalence of IPV among women, and exacerbated IPV-related health impacts, including injuries, depression and posttraumatic stress disorder (PTSD). [21]There have been significant consequences of physical and psychological violence against women during the COVID-19 pandemic. [17] Given the seriousness to the problem, the UN Secretary General Antonio Guterres called for measures to address this rising of domestic violence directed toward girls and women linked to lockdowns imposed by governments responding to the pandemic. [22]

Timor-Leste (TL) is a country with significant rates of D-GBV with One in three Timorese women have experienced domestic violence [23] Although the prevalence is low at 0.30%, evidence found that an evolution toward an HIV/AIDS epidemic is starting in TL. [24] Women living with HIV, female sex workers, men who have sex with men and transgender women have difficulties negotiating for safer sex and accessing prevention commodities. [25] With the incidence to 486 per 100,000 TL rank as the second highest TB incidence rate in the South-East Asia region. [26] The first case of COVID-19 on 1 January 2020 and by the 30 of June 2022, the country counted 23,433 COVID-19 cases, with 753 persons were hospitalised and 133 died due to COVID-19. [27] From April 2020 until the end of the pandemic, the country imposed strict lockdowns. [28] The strict lockdowns were important as they formed a backdrop to the relatively low case numbers in 2020 and curbed case incidence in 2021. [29] [30] However, the lockdown also caused the disruption of work, school, health services provision, slowed down the economic and reduced the country’s capacity to achieve the SDGs. [31] [32] [33] [34] Caritas Australia reported an increase of GBV due to COVID-19 and believed that factors such as financial stress, close confinement of families, and isolation from support networks contributed to the increase of violence. [35]

Despite, there is a well-established link between the COVID-19 pandemic and the increased number of cases of GBV, tuberculosis, and HIV infections. However, to our knowledge, there are limited numbers of research examining the occurrence of D-GBV against girls and women living with tuberculosis and HIV/AIDS during the pandemic in developing countries. The Ministry of Health (MOH) and the Global Fund (GF) Timor Leste commissioned this study to understand whether women living with these two diseases experienced D-GBV during the lockdowns.

## METHOD

This is a qualitative phenomenology study utilizing purposive sampling technic to enroll participants and explore the phenomenon of domestic gender-based violence that occurred in women and girls living with tuberculosis and HIV/AIDS during the COVID-19 pandemic in TL. The study was conducted in eight municipalities from early October 2022 to end of February 2023. It considered eight independent variables – physical violence, verbal violence, phycological violence, sexual violence, sexual harassment, social-economic violence, intimate partner violence (IPV), and stigma and discrimination – to identify the occurrence of GBV, see Table-2 (become-table-1). The study included 42 in-depth interviews (IDIs) with 19 HIV participants and 23 TB participants, and three focus group discussions (FGDs), see-**Error! Reference source not found**.2.

## SETTING

This study was conducted in eight municipalities, including Ainaro, Bobonaro, Covalima, Ermera, Liquiça, Manufahi, RAEOA and Dili (figure-1). Timor-Leste is a small, half island country of 15,007 km^2^ divided into 13 municipalities and one autonomous region (Oecuse), 67 administrative posts and 452 villages. It is is a peaceful country, democratic nation with a population of 1,340,434. [36] Poverty levels remain high despite progress made in improving the standards of living. The human capital index in 2020 stood at 0.45, lower than countries from East Asia and the Pacific of 0.59. [37] Health has been a priority since Timor-Leste gained independence, as clearly demonstrated in the RDTL’s Constitution, where article 57 guarantees the fundamental right of each citizen to access free health care. [38] Timor-Leste’s referral system includes three levels: a national tertiary hospital, five referral hospitals and community health centres (CHCs), and health posts (HPs). At the municipal level, the delivery of the primary health care (PHC) is done through the CHCs, HPs, integrated community health services (SISCa), and the *Saúde na Família* (SnF) network. Currently, there are 6 hospitals, 72 CHCs and 329 HPs, directly linked to the SISCa and SnF posts operating across the territory. [39] Health Indicators have improved substantially since 2002 as reported in the Timor-Leste Demographic Health Survey (TLDHS) 2016. [23]

## DATA COLLECTION

The research team took a series of steps for the data collection, including study site visits and enrolment and interview of the study participants.

a. The research team compose of Co-Investigator (Co-I) and Research Assistant (RA), visited the municipalities where they met with the President of the Municipal Authority and the Director of the MHS, and handed a letter from the principal investigator (PI) outlining information on and purpose of the study,
b. The research team purposely identified CHCs for the study where they met with the head and the TB and HIV units to introduce the study and request permission for conducting the interviews.
c. The research utilizes developed procedures for selecting, enrolling and interviewing study Participants was described in table-3.

## DATA ANALYSIS

Data analysis (coding) for overall interview results was performed by an independent consultant utilizing NVIVO version 12.1 pro, year 2020, by identifying codes, themes, and categories. In addition, each co-investigator was tasked to transcribe and manually code the interview results. The combination of deductive and inductive thematic analysis approach was adopted to analyse the information obtained from the interviews. This helps researchers identifying information according to pre-determined themes, as well as developing new themes emerges from the data.

To ensure the validity and reliability of the results, the data was read, re-read, and themes (repeated patterns of meaning) were generated from the data. The six steps from Braun and Clarke (2006) were adopted during the process of data analysing and data coding. Those six steps were: (a) familiarizing yourself with your data, (b) generating initial codes, (c) searching for themes, (d) reviewing themes, (e) defining and naming themes, and (f) producing the report. [40]

The results generated by the independent consultant with NVIVO software were presented to the forum of co-investigators for clarification, checking for similarities, and ensuring the consistency. The principal investigator compared both coding results (the coding results from NVIVO software vs manual coding) to derive the final conclusions for each pre-determined theme and category. If there was discrepancy found between two codes, both Independent Consultant and Co-Investigator will be called to clarify and agree on the final meaning and concept. Findings from the in-depth interviews, FGDs and field note observations were further triangulated to develop the final, comprehensive results for each pre-determined theme.

## ETHICAL CONSIDERATION

The study obtained the ethics approval from National Health Institute-Health Research Ethics Committee, MOH-Timor Leste, with Ethic Approval number : 229/MS-INS/GDE/X/2022).

Each participant was informed about the aim of the study and there would be no consequences if they withdraw from the study without giving any reason. All participant signed and returned a written informed consent to research team before interview which took 30–50 minutes, and the interview was recorded with the consent of the participants. Identification letter and number based on diseases, participants, and municipalities (e.g. TB-Ainaro-1, HIV-Dili-2) was used for confidentiality purposes.

## RESULTS

Total of 42 IDIs and three FGDs were conducted with girls and women living with HIV and tuberculosis. IDIs were carried out for 19 HIV participants and 23 with TB participants. One FGD with HIV participants was conducted in Dili, and two FGDs with TB participants were conducted in Liquiça and Covalima. Most of the participants enrolled in this assessment were women, married and above 20 years old, living in rural areas of Timor-Leste. The findings were categorized into several themes (see Fig. 2) which are used to structure this section of the paper.

### 1) MAGNITUDE OF D-GBV

Domestic GBV were prevalent all municipalities, where all municipalities reported more than three types of violence. For HIV participants, psychological, socio-economic, verbal, and physical violence were reported the most from five municipalities (**Error! Reference source not found**.). Similarly, female suffered tuberculosis from all seven municipalities reported on psychological, socio-economic, verbal, and physical violence (**Error! Reference source not found**.). The COVID-19 pandemic exacerbated D-GBV and impeded girls and women living with HIV and TB to apply coping mechanisms in dealing with the violence

### 2) VULNERABILITY For D-GBV

This study identified a “triple vulnerability” for female HIV and TB patients experienced GBV during the COVID-19 pandemic.

The first vulnerability is associated of a being a girl or a woman living in a patriarchal society in Timor-Leste with a strong culture /tradition that requires women to play traditional household roles in the society and at the household level, making them prone to D-GBV. A failure to perform these roles may result in domestic violence against them.

A married woman interviewed in this study argued that continuous experience of violence from her husband caused her to get infected with tuberculosis : “*My problem at home that, my husband beat me every day, kicked my back, punched, slapped that’s the problem, not loving us want us to die, curse us to die, that’s why I got sick like this,…because of…building a secret house in the afternoon, said we have to go back, when he doesn’t want to go back, he hit me on my back, that’s why I got sick like this, we argue we fought, he hit me with hand*.*” TB-FGD Covalima*

Marrying at a young age without economic stability makes the young woman more vulnerable to domestic violence: “*My husband is the one who was beating me. We married at a young age and stayed with my mother-in-law, and because we didn’t have money, we often had conflict and he often beat me*.*”* (HIV-Dili-6)

Although she understands or is knowledgeable, the woman is not allowed to convey her opinion openly:*” Yes, our tradition, when our husband/elder spoke small or big quiet, women not to talk back (not to talk much), you know, you women also know tradition, so women not talking*.*” FGD-TB Covalima*.

The second vulnerability is related being a female and suffered HIV and TB living in Timor Leste. TB and HIV diseases reduces the physical and psychological capacities to perform routine household roles, which in turn increases the incidence of D-GBV.

Female HIV and TB patients expressed that due to weakness and tiredness they often failed to perform regular household work in which triggers the violence: “*Yes, I often got scolded, yelled at, and screamed at. They told me to leave home and often blamed me even when I was doing housework. Though I am sick, but they kept asking me to work, and if didn’t work, I would not eat*.*"(HIV-Dili1)*

A married woman often carries an even higher burden when falling sick. She needs to continue perform her responsibility as a mother and take care of the household. Very often, family members do not understand or refuse to understand the sick woman’s conditions, which in turn causes violence.:*"Because they don’t understand my condition, I don’t do my responsibilities because I’am sick (not normal)", patient sad and crying*.*” (TB-Manufahi-1)*

The sick married woman who stays together with family, experiences violence not only from her husband, but also from other family members: “*We live together with in-laws, during the time I was sick my mother-in-law always yelled at me, saying because of sickness you don’t want to work, there is medication always drink it and do work, if you don’t want to work who’s going to feed you and the kids*.*” FGD-TB Liquica)*

The third vulnerability is related to being a girl or woman with HIV/TB staying at home during the COVID-19 lockdown in Timor Leste, will further increased the number of D-GBV cases. The factors that mostly triggered the violence were income loss and long periods of time spent together with the perpetrator at home.

“*My husband worked at a store in Dili, but because of the COVID-19 outbreak, my husband didn’t work and only stayed at home. I was also not working, just looking after my children, and we lived with my parents-in-law, when I was sick my parents-in-law took care of me. Without money, we sometimes got angry. When there is no work, sometimes it makes us angry at each other…*, ***When my husband was angry, he would beat and swear at me. Just because of the COVID-19 came, our products were not sold***, *there was also no money, which caused us problem*. ***I was stressed because I didn’t have any money and my disease was making me easily tired when doing heavy work***". (TB-FGD-Bobonaro)

### 3) THE PERPETRATOR

The perpetrator of physical violence were mostly husbands (intimate partner violence), as pointed out by an HIV patient interviewed in Dili. “*My husband is the one who was beating me. We married at a young age and stayed with my mother-in-law, and because we didn’t have money, we often had conflict and he often beat me*.*”* (HIV-Dili-6)

Verbal and psychological violence was committed by husbands, family members and community, as illustrated by a woman living with HIV and TB :*"I got so many insults from my brother and sister-in-law; they kicked me out of the home*.*”* (HIV-Dili-4);

A girl living with tuberculosis said she did not get any physical violence from her family, but she often got verbal violence from her neighbour.*"But I felt sad and worried with my neighbours because they sometimes insulted me by asking me how come you and your family had this TB, your mother just recovered and now you have it. It was a big concern for me because I am a student who is still studying at high school, and I am also afraid that my friends will stay away from me and also become enemies with me due to the disease that I have*.*”* (TB -Liquiça-1).

### 4) TYPES OF DOMESTIC VIOLENCE AND GENDER-BASED VIOLENCE IDENTIFIED

All types of violence were reported from both HIV and TB participants from all municipalities, with psychological violence was the most reported and sexual violence as the least reported during the interview. The acts of violence for each type of violence is listed in the table-4, and below are the original quotes on three types of violence :

Psychological Violence : “*During COVID-19, family isolate me at home and not letting me out in the community or contact with other people because they were afraid of me get infected with COVID-19 and causing a high risk to my health with the reason I have a TB disease, it can cause complications, and at times my colleagues asked me to participate in extra activities I cannot participate and I feel stressed or a lot of thinking start appearing in my mind (thinking a lot). TB-Liquiça-1-Girl*”

Socio-economic Violence (Loss of Income) : “*My family lives of farming, my husband divorced me, and I felt sad because I do not have husband anymore and the children keep asking me where their dad is. We don’t have money and there is no money to buy rice. During COVID-19, we didn’t have money and there was no food. I sold out all my gold necklaces to get money to buy rice. I didn’t have any strength to work and find a job and I only stayed at home. Sometimes my dad gave me $20 to buy my necessities but didn’t give me money to manage. HIV-Dili-4”*

Sexual Violence (Force) : “*But from my husband, he often forces me. I don’t like being forced. If we have desire then we can do, if not then I don’t want. But as a wife you have to do your responsibility*…, *I just accept it, it was my decision to marry, so if my husband forces me to have sex, I have to accept it. TB-Ermera-1”*

### 5) CAUSES OF THE VIOLENCE

Various causes of violence identified from the interviews were including: family interferences, coming back home late, jealousy and denial, alcohol drinking, economic factors, kids’ school fees, no sharing of responsibilities, sickness, tradition/cultural issues, failure to perform household work, miss-communications, and engaging in an argument. Below is the quote by one of TB respondent on the economic factor and family responsibility that triggers the violence. “*I want to share the responsibility of looking after the children’s school and supporting my children’s studies. When I told him about the children’s tuition fees, he answered with anger (swearing) and told me to stop sending the children to school. I’m very sad. For me school is very important. I didn’t go to school, so I want my children go to school. TB-Manufahi-1”*

### 6). COPING MECHANISMS

Girls and women living with TB and HIV reported using various coping mechanisms or strategies to deal with domestic violence and gender-based violence. These coping mechanisms can be categorized into two groups of actions:

1. seeking external support or avoidance with acts of seeking assistance from formal institutions such as military and police force, nuns, and NGO, seeking protection and assistance from parents and family, going out, using avoidance, divorcing, being economically independent, and receiving/seeking support from neighbours, *Quotes:* “*I inform brothers FDTL (military) and UPF (police) that the sisters (nuns) will resolve. HIV Bobonaro2”* and
2. staying and facing the perpetrator at home with acts of staying quiet and patient, paying for support and protection, Praying, Ignoring, perceived as normal household roles, Acceptance and praying. Quotes: Stay Home and facing Perpetrator (Acceptance and praying):*"I accept this suffering because of my children’s studies. My children are my strength, so I have to do my best for everything though I have to be hurt. The most important is that they are cleaver and succeed. So, I don’t want my husband to use violence with my children, just use violence with me. I continue to pray to God to give me strength, a long life, so that I can be patient to accept this violence and forgive my husband. I want to recover from TB and get well with the treatment. TB-Ainaro1*

### 7) STIGMA AND DISCRIMINATION

Stigma and Discrimination were widely experienced by participants living with HIV and tuberculosis. Various forms of discrimination acts committed against female HIV and TB patients were including: discriminative words towards the children; neighbours band visiting their house, utensils separation, distancing, avoiding contact, difficult to get information or to access health care, banned to work, social/race discrimination, and clothes separation. Below are two of the original quotes from a female HIV and a female TB participant:

” *it’s a close family living together, who talks like this…the sick (bad disease) people’s clothes don’t wash together…otherwise the disease can infect us, and told the kids to not get close to her, she had a bad disease. HIV-Bobonaro-1*

“*my neighbour said don’t get close to me because I have TB disease and I could transmit my disease. They often talk about me and talk about my disease. TB-FGD Liquica*

## DISCUSSION AND CONCLUSION

The Magnitude of D-GBV During COVID-19 Pandemic:

The epidemic of domestic violence against women worldwide greatly intensified during the COVID-19 pandemic. [19] The COVID-19 pandemic and its measures such as quarantines and social distancing increased women’s exposure to violence as confinement in physical spaces along with economic and health shocks have increased household stress levels. [41] Globally, there are increasing number of studies and reports documented the impact of COVID-19 on the D-GBV. [17] [20] [21] [42] [43] [44] However, most of them were focusing on examining the violence against healthy girls and women during disease pandemic. Our study contributes qualitative information on the D-GBV committed against female HIV and tuberculosis patients during COVID-19 Pandemic.

Although this is a qualitative study with female HIV/TB patients only, the findings have amplified the qualitative information on the magnitude of D-GBV in TL. Before the COVID-19 pandemic, reports highlighted high prevalence of GBV among healthy women in TL.

[23] [45] Our study identified that girls and women living with HIV and TB suffered various forms of D-GBV, including psychological, verbal, socio-economic, physical, and sexual violence, as well as stigma and discrimination during the COVID-19 pandemic.

Findings from this qualitative study suggest the need to pay more attention to female HIV and TB patients in order to help them survive the disease(s) and protect them from GBV, especially during disease pandemic and future humanitarian crisis in Timor-Leste. Evidences from previous studies reveal that although many of the HIV clinical features in women are similar to those in men, there remain significant sex-based differences in the disease. [46] Globally, data from before the COVID-19 pandemic shows that TB kills more women than any other infectious disease, and more women die annually of TB than of all causes of maternal mortality combined. [47] Assessing these female conditions and choosing appropriate diagnostic and treatment procedures by clinician trained in both infectious diseases and GBV will better help the victims survive those infectious diseases and the impact from D-GBV.

### Vulnerability for D-GBV

The first vulnerability associated with patriarchal society in TL is quite similar to the experience for other developing countries, especially countries in the Asia-Pacific Region. Previous research reveal South Asian women’s experiences of abuse operate along a continuum of cultural and social values and beliefs. In their social structure women shall play role as good wives, to maintain harmony, and to take care of the family at home, while men be the head and provider for the family. [48] In Pakistan, patriarchal values are rooted in Pakistani society which regulates the subordinated position of women. [49] GBV is arguably grounded on an inequality of power and is often committed for various reasons from power imbalance, cultural, war and migrant status triggers men to commit violence against women. [3]

The second vulnerability linked to HIV and TB infections that reduces female physical and psychological capacities to perform activities including household work is well proven in medical literature. Similar to men, female with TB and HIV infectious often experience symptoms and signs that reduce their physical and psychological capacities. These symptom and signs are including weakness, shortness of breath, dizziness, tiredness, loss of appetite, loss of weight, cough, frequent low-grade fever, and night sweats, and nauseous. [50] [51] [52] If the violence continues, those women may suffer another form of the diseases as a consequence from the violence. Previous studies identified a number of impacts that resulted from violence, including reproductive disorders such as menstrual irregularities and disorders in the process of pregnancy, and mental disorders such as the emergence of anxiety, fear, fatigue, and stress, even frequently have an impact on eating and sleep disorders, and serious physical and economic. [17] [42]

The third vulnerability linked to female HIV/TB staying at home during COVID-19 lockdown will further increased the number of D-GBV. This is true as the COVID-19 pandemic and its lockdown measure deepened economic and social stress, caused substantial increases of physical and psychological violence against women, especially IPV among women and exacerbated IPV-related health impacts, including injury, depression, and PTSD. [17] [20] [21]

### Perpetrator and Trigger of D-GBV

The acts D-GBV identified in this study was committed mostly by husband and categorized as intimate partner violence (IPV). This findings conform to the WHO, 2012 report that claim IPV as one of the most common forms of violence against women and argues that it occurs in all settings and among all socioeconomic, religious, and cultural groups. [53] IPV refers to any behaviour within an intimate relationship that causes physical, psychological, sexual harm and *controlling* behaviours. [53] [54] In this study, we identified a number of similar acts to those reported by WHO above, including husband controlling behaviour. Kishor and Johnson (2004) identified six main controlling behaviours that link to IPV, such as: (1) husband becoming jealous or angry if the wife talks with another man, (2) husband frequently accusing her of being unfaithful, (3) husband not permitting her to meet girlfriends, (4) husband limiting her contact with her family, (5) husband insisting on knowing where she is all the time, and (6) husband not trusting her with money. [55] In this study, we identified jealously, limiting contact with family, accusing unfaithful, and not trusting with the money as means of husband controlling behaviour against the victims.

This study found number of conditions that triggered D-GBV against female TB and HIV patients during the pandemic and lockdown period. Those conditions included family interferences, coming back home late, jealousy and denial, alcohol drinking, economic factors, no sharing of responsibilities, sickness, tradition & culture, failure to perform household work, and miscommunication. The similar conditions were well documented elsewhere from previous studies. The disruption of livelihoods, reducing the ability to earn a living to meet basic needs, and coupled with more time spent in close contact in cramped conditions, causing additional stress which in turn triggers violence. [43] [44] Other scholar argue that the power imbalance, cultural, war and migrant status, trigger men to commit violence against women.. [3] Russo, 2006 argues that to understand, predict, and prevent gender-based violence will require a complex and comprehensive approach that intervenes at individual, interpersonal, and structural levels and that is responsive to cultural difference. [54]

### Stigma and discrimination

Stigma and discrimination were widely experienced by participants living with HIV and tuberculosis from this study. It is internationally recognized that GBV is associated with stigma and shame and has negative consequences on the victims/survivors. Stigma and the associated emotion of shame combine to become a powerful form of social control. [54] According to Goffman, 1963 stigma is “an attribute that is deeply discrediting” that reduces someone “from a whole and usual person to a tainted, discounted one". [56] Shame has been identified as a factor in inhibiting women from disclosing their experiences of violence to others and from seeking help. [57] Shame is a complex emotion and often discussed with reluctance; these feelings are usually incapacitating, and unbearable shame is an overwhelmingly negative emotion. Feelings of shame make someone feel insignificant and inferior with a wish to be invisible and not be noticed. [58] Indeed, as experienced from this study some of the participants bear negative consequences resulted from the stigma and shame. As identified in this study, shame is more associated with HIV diseases and Sexual Violence which caused the victim to hide their identity and refuse talk openly about their experience to the outsiders.

### Coping Mechanisms to Endure D-GBV

Despite the fact that girls and women suffer from the diseases and experience regular D-GBV, they have developed various mechanisms to cope with the consequences of violence. Two main categories of coping mechanisms were adopted by the victims to survive both their disease and the violence. The first category includes acts of seeking external support, going out, adopting avoidance, and seeking family protection. The second category includes acts of staying and praying, acceptance, crying, and remaining quiet. The lockdown has impacted heavily the women from first category. According to Elisabeth and colleagues, 2022, the lockdowns disrupted the social and protective networks used by women to have contact with family and friends who provide support and protection from the violence perpetrated by a partner. It restricted women from accessing sexual and reproductive health services, psychological support, and essential support services such as hotlines, crisis centres, shelters, legal aid, protection, and counselling. [43]

### Study Limitations and Strength

This is the first qualitative study conducted in Timor-Leste to assess GBV against female HIV and TB patients during the COVID-19 pandemic. Although the findings are novel and provide an important contribution to the global scientific community, the interpretation of the findings needs to consider three of the main limitations associated with the nature of the methodology chosen for this study.

First, since this nature of qualitative study is not design to quantify and generalize the finding, therefore the results cannot be generalized to the whole country. Creswell, 2018, define qualitative research as an inquire process of understanding based on distinct methodological traditions of that explore a social and human problems. It builds a complex, holistic picture, analyses words, reports detailed views of informants, and conducts the study in natural settings. [59]

Second, the selection of the participants was by nature biased. With reference to the time, complexity, accessibility, and confidentiality attach to people living with GBV, HIV, and TB, a sample size between six and nine participants for each municipality was determined. A. Moser and I. Korstjens (2018) argue that sample size in qualitative research can be defined on the basis of “the rule of the thumb". [60]

Third, interview biases: the interview were conducted by three researchers with different levels of seniority, which may have derived to some interview biases, especially for interviews conducted by counsellors and research assistants in local languages. On the other hand, talking in local languages helped participants to better express their feelings and opinion. In addition, interviews conducted by counsellors who known the patients create a better and friendlier environment for participants to express their opinion, experiences and feelings when answering the research questions.

## Conclusion

This was the first qualitative study conducted to identify D-GBV against girls and women living HIV and tuberculosis in Timor-Leste. its findings have amplified the qualitative information on the magnitude of D-GBV in Timor-Leste. The psychological, verbal, socio-economic, physical, and sexual violence, as well as stigma and discrimination were prevalent in all study municipalities. Female TB and HIV living in Timor Leste were facing triple vulnerable situation for D-GBV during COVID-19 pandemic. The victim of D-GBV applied two main categories of coping mechanisms were adopted by the victims to survive both the diseases and the violence. The study recommends to sensitize all organizations working in health and on gender and on ending violence against women (EVAW) on the magnitude of D-GBV. Train doctors, nurses and other health personnel working in the areas of tuberculosis and HIV control on D-GBV to assist the survivors when needed.

## Data Availability

All data produced in the present study are available upon reasonable request to the authors

## AUTHOR CONTRIBUTIONS

Conceptualization: Nelson Martins, Domingos Soares, Caetano Gusmao, Jorge Martins, Valente da Silva

Formal Analysis: Marcelo Mali, Domingos Soares, Maria Nunes, Nelson Martins

Investigation: Domingos Soares, Caetano Gusmao, Valente Da Silva, Maria Nunes, Diana Valadares, Laura Abrantes, Suzie Marcal

Methodology: Nelson Martins, Domingos Soares, Caetano Gusmao, Valente Da Silva

Project Administration: Luis Alves and Jorge Martins

Writing Original Draft : Nelson Martins

Writing – review and editing: Nelson Martins and Domingos Soares

## ACKNOWLEDGMENT

To Research assistants, Counsellors, and all individuals and organisations that indirectly provided support during the conception, proposal development, presentation, field data collection and report writing.

